# Household and Maternal Characteristics Associated with Malaria Prevalence Among Children Under Five Years in Ghana: Evidence from the 2019 Malaria Indicator Survey

**DOI:** 10.64898/2026.08.23.26361158

**Authors:** Eric Ohemeng

## Abstract

**Background:** Malaria remains a major cause of morbidity and mortality among children under five years in Ghana. This study examined household, maternal, and child characteristics associated with malaria prevalence.

**Methods:** Data were drawn from the 2019 Ghana Malaria Indicator Survey. The analytic sample comprised 2,867 children with valid malaria rapid diagnostic test results. Analyses accounted for the survey’s multistage cluster design, including primary sampling units, strata, and sampling weights. Survey-weighted descriptive statistics, Rao-Scott corrected chi-square tests, and survey-weighted logistic regression with a quasibinomial link were used.

**Results:** Overall, 22.9% of children tested positive for malaria. At the bivariate level, child age, number of children under five, household wealth, residence, region, household net usage, electricity, television ownership, maternal education, household size, and anaemia level were significantly associated with malaria status (p < 0.05). In the fully adjusted model, children in the poorest and poorer households had higher odds of testing positive than those in the richest households (AOR = 4.13 and 3.13, respectively). Rural children had higher odds than urban children (AOR = 2.36). The highest regional odds were observed in the Eastern Region (AOR = 15.3) compared with Greater Accra. Older children and those with severe anaemia had higher odds of testing positive (AOR = 4.17 and 24.5, respectively). Maternal education significantly interacted with household wealth and household net usage.

**Conclusion:** Household wealth, region, residence, maternal education, child age, and anaemia level were important correlates of malaria prevalence. Findings support interventions addressing socioeconomic and regional inequalities in childhood malaria.

## 1. Introduction

Malaria continues to be a major global public health problem among children under five years, despite decades of control and elimination efforts (Dao et al., 2021). The World Health

Organization’s 2021 World Malaria Report estimated 241 million malaria cases globally in 2020, an increase of 19 million cases from 2019, with the WHO African Region accounting for 95% of global cases (World Health Organization, 2021a). Children under five years bear a disproportionate share of this burden. Although the proportion of malaria deaths among under-five children declined from 87% in 2000 to 77% in 2020, this age group still accounts for the majority of malaria-related deaths in Africa (World Health Organization, 2021b).

In Ghana, malaria remains the leading cause of outpatient department visits covered by the National Health Insurance Scheme, accounting for approximately ten million visits annually, about four percent of which are among children under five years (Ejigu & Wencheko, 2021). Although national interventions such as the distribution of insecticide-treated nets (ITNs) and the 2015 Global Technical Strategy for Malaria have reduced malaria mortality, morbidity and outpatient caseloads among children remain high (Afoakwah et al., 2018; Okyere, 2021).

Much of the existing literature on malaria control in Ghana and elsewhere in sub-Saharan Africa has emphasized macro-level interventions, such as vector control and health system strengthening, while paying comparatively less attention to the household- and maternal-level characteristics that shape a child’s risk of infection. Yet these characteristics, including household wealth, maternal education, place and region of residence, and use of preventive measures, are theorized to influence a household’s capacity to produce and protect child health (Becker, 1965; Grossman, 1972; Mosley & Chen, 1984; Solar & Irwin, 2010). Evidence on the direction and strength of these associations, however, remains mixed across settings. Maternal education, for example, has been linked to a lower risk of childhood malaria in Uganda and Ghana (Masuda, 2020; Ejigu & Wencheko, 2021), while findings on household wealth (Filmer, 2005; Yusuf et al., 2010; Bayode & Siegmund, 2022) and place of residence (Sultana et al., 2017; Iqbal et al., 2016; Savi et al., 2021) are inconsistent.

Few recent Ghanaian studies have used a nationally representative, malaria-specific survey to examine how household, maternal, and child characteristics jointly relate to malaria prevalence among children under five years, and even fewer have examined how malaria prevalence relates to concurrent child anaemia status in this population. This study addresses that gap using data from the 2019 Ghana Malaria Indicator Survey (GMIS). Specifically, the study examines (i) the relationship between household characteristics and malaria prevalence among children under five years, (ii) the association between malaria preventive measures and malaria infection among children under five years, and (iii) the extent to which maternal education and child anaemia status relate to malaria prevalence among children under five years.

## 2. Conceptual Framework

This study draws on the household production-of-health framework, which conceives of child health as an output that households produce by combining structural resources with time, knowledge, and preventive behaviors (Becker, 1965; Grossman, 1972). Mosley and Chen’s (1984) analytical framework for child survival operationalizes this idea for low- and middle-income settings by distinguishing household-level determinants from the intermediate behavioral determinants through which they act on a child’s health outcome, while the World Health Organization’s social determinants of health framework (Solar & Irwin, 2010) similarly emphasizes that structural position shapes health through intermediary factors such as material circumstances, behaviors, and access to services.

Adapting these frameworks to childhood malaria (Figure 1), household size, place of residence, electricity, region of residence, sex of the household head, number of children under five in the household, and household wealth status are treated as household characteristics that shape a household’s capacity to produce health and its exposure to malaria risk. These household characteristics act on malaria prevalence both directly and through a set of intermediate variables — whether the dwelling was sprayed, the number of mosquito nets owned, and the number of children who slept under an insecticide-treated net (ITN) — which represent the preventive behaviors and resources households draw on to protect children from infection. Sex of the child, age of the child, maternal educational level, and the child’s anaemia level are included as child and maternal characteristics that act on malaria prevalence both directly and by shaping household preventive behavior. This study’s finding that mother’s education interacts significantly with both household wealth and household net usage (Section 4.4) suggests that, consistent with Solar and Irwin (2010) and Mosley and Chen (1984), household and maternal characteristics can moderate the effectiveness of intermediate variables rather than operating on malaria prevalence only through them.

**Figure 1.**
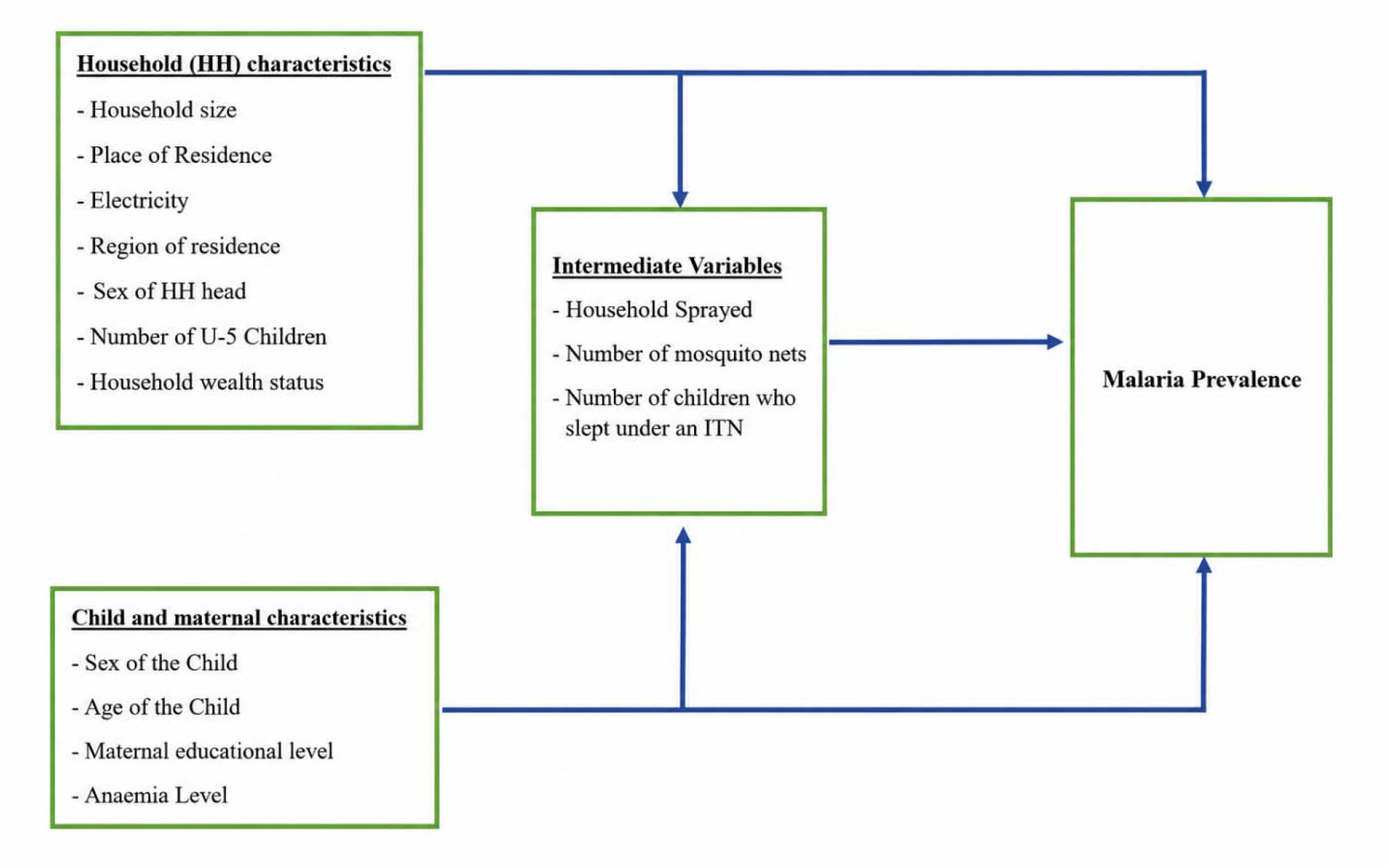
Conceptual framework linking household characteristics, intermediate variables, and child and maternal characteristics to malaria prevalence among children under five years.

## 3. Methods

### 3.1 Study design and data source

This study used secondary data from the 2019 Ghana Malaria Indicator Survey (GMIS), a nationally representative, cross-sectional household survey implemented by the Ghana Statistical Service (GSS), the Ghana Health Service, and the National Malaria Control Programme, with funding from the Global Fund and the Government of Ghana. The GMIS follows the Demographic and Health Surveys (DHS) Program protocol, adapted to include malaria-specific content. As this study used secondary, de-identified survey data, additional ethical approval was not required beyond the approvals obtained for the original survey by the Ghana Statistical Service and ICF (Ghana Statistical Service & ICF, 2020).

### 3.2 Sampling and study sample

The 2019 GMIS used a stratified two-stage sample design based on the 2010 Ghana Population and Housing Census sampling frame. Each of Ghana’s ten regions (at the time of the survey) was stratified into urban and rural areas, yielding 20 sampling strata. In the first stage, enumeration areas were selected with probability proportional to size; in the second stage, households were randomly selected within each cluster. The analytic sample for this study comprised all children under five years with a valid malaria rapid diagnostic test (RDT) result recorded in the household member file (n = 2,867). To account for the survey’s multistage cluster design, all descriptive and inferential analyses incorporated the primary sampling unit, sampling strata, and normalized household sampling weight.

### 3.3 Measures

The outcome variable, malaria prevalence, was defined as a positive rapid diagnostic test result (yes/no). Household-level characteristics included household size (<6, 6–9, >9 members), number of children under five in the household (0–1, 2–3, >3), sex of the household head, household wealth status (DHS wealth index quintiles, poorest to richest), place of residence (urban/rural), region, and household electricity. Intermediate (preventive-behavior) variables included the number of mosquito nets owned (0 to 7 or more), the number of children under five who slept under a net the previous night (no child, 1–2 children, more than 2 children), and whether the dwelling had been sprayed against mosquitoes in the previous 12 months. Child-level characteristics included age in months (<24, 24–48, >48), sex, and anaemia level (not anemic, mild, moderate, severe), measured by hemoglobin concentration adjusted for altitude. Maternal education was categorized as none, primary, secondary, bachelor’s, or higher.

### 3.4 Statistical analysis

Analyses were conducted in R using the survey package to account for the complex sample design (Lumley, 2004). A survey design object was defined using the primary sampling unit as the cluster identifier, the sample strata variable, and the normalized household sampling weight. Weighted descriptive statistics (svymean) summarized the distribution of household, maternal, and child characteristics, and bivariate associations with malaria status were tested using Rao– Scott second-order corrected Pearson chi-square tests (svychisq), which adjust conventional chi-square statistics for the design effect of cluster sampling. Multivariate analysis used survey-weighted logistic regression (svyglm) with a quasibinomial link to accommodate the design-based dispersion adjustment. Three nested models were estimated: Model 1 included household characteristics only (household size, number of children under five, sex of household head, wealth, residence, region, electricity); Model 2 added mother’s education and preventive-behavior variables (dwelling spraying, number of nets owned, children’s net usage); and Model 3 (the final model) added child-level characteristics (child’s age, child’s sex, and anaemia level). Two-way interaction terms between mother’s education and household wealth, and between mother’s education and children’s net usage, were each tested against the final model using design-based Wald F-tests (regTermTest). Statistical significance was assessed at α = 0.05, and adjusted odds ratios (AOR) with 95% confidence intervals are reported throughout.

For a child i with sampling weight w□, let π□ denote the survey-weighted probability of a positive malaria test. The Horvitz–Thompson survey-weighted mean used for the descriptive statistics in Table 1 and Table 2 is given by:

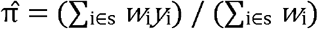

**Table 1.** Weighted characteristics of children under five years with a valid malaria test result, 2019 Ghana Malaria Indicator Survey (n = 2,867). Estimates and standard errors account for the survey’s cluster sampling, stratification, and sampling weights. Source: Computed from GMIS 2019 using the R survey package.

| Characteristic | Weighted % | SE |
| --- | --- | --- |
| **Malaria test result** |  |  |
| Negative | 77.1 | 0.017 |
| Positive | 22.9 | 0.017 |
| **Child's age** |  |  |
| < 24 months | 34.4 | 0.011 |
| 24–48 months | 45.6 | 0.013 |
| > 48 months | 20.0 | 0.008 |
| **Sex of child** |  |  |
| Male | 50.5 | 0.010 |
| Female | 49.5 | 0.010 |
| **Number of children under five in household** |  |  |
| 0–1 | 41.6 | 0.016 |
| 2–3 | 51.0 | 0.012 |
| > 3 | 7.4 | 0.011 |
| **Household size** |  |  |
| < 6 members | 49.7 | 0.017 |
| 6–9 members | 37.6 | 0.011 |
| > 9 members | 12.7 | 0.014 |
| **Sex of household head** |  |  |
| Male | 68.4 | 0.018 |
| Female | 31.6 | 0.018 |
| **Household wealth status** |  |  |
| Poorest | 24.1 | 0.023 |
| Poorer | 22.1 | 0.015 |
| Middle | 21.0 | 0.016 |
| Richer | 18.1 | 0.016 |
| Richest | 14.8 | 0.014 |
| **Place of residence** |  |  |
| Urban | 40.5 | 0.024 |
| Rural | 59.5 | 0.024 |
| **Mother's education** |  |  |
| None | 22.8 | 0.018 |
| Primary | 21.8 | 0.016 |
| Secondary | 37.8 | 0.019 |
| Bachelor's | 12.4 | 0.011 |
| Higher | 5.2 | 0.007 |
| **Mosquito net ownership (any ITN)** |  |  |
| No | 0.2 | 0.001 |
| Yes | 99.8 | 0.001 |
| **Number of mosquito nets owned** |  |  |
| 0 nets | 14.2 | 0.013 |
| 1 net | 17.2 | 0.014 |
| 2 nets | 23.2 | 0.013 |
| 3 nets | 19.6 | 0.014 |
| 4 nets | 12.8 | 0.011 |
| 5 nets | 5.7 | 0.008 |
| 6 nets | 3.5 | 0.006 |
| 7+ nets | 3.9 | 0.007 |
| **Children's mosquito net usage**<br>**(household level)** |  |  |
| No | 25.8 | 0.016 |
| All children | 47.8 | 0.018 |
| Some children | 12.1 | 0.013 |
| No net in household | 14.3 | 0.011 |
| **Number of children under 5 who slept under a net** |  |  |
| No child | 40.5 | 0.017 |
| 1–2 children | 52.8 | 0.017 |
| > 2 children | 6.7 | 0.010 |
| **Dwelling sprayed (12 months)** |  |  |
| No | 89.7 | 0.027 |
| Yes | 10.3 | 0.027 |
| **Household electricity** |  |  |
| No | 21.5 | 0.026 |
| Yes | 78.5 | 0.026 |
| **Household television** |  |  |
| No | 37.8 | 0.020 |
| Yes | 62.2 | 0.020 |
| **Child's anaemia level** |  |  |
| Not anemic | 45.2 | 0.021 |
| Mild | 26.7 | 0.017 |
| Moderate | 26.7 | 0.016 |
| Severe | 1.4 | 0.004 |

**Table 2.** Weighted malaria prevalence (95% confidence interval) by household, maternal, and child characteristic, with Rao–Scott second-order corrected Pearson chi-square p-values. *p < 0.05. Source: Computed from GMIS 2019 using the R survey package.

| Characteristic | Category | Malaria prevalence %<br>(95% CI) | p-value |
| --- | --- | --- | --- |
| **Household size** |  |  | * |
|  | < 6 members | 19.9 (16.7–23.1) | 0.021* |
|  | 6–9 members | 24.8 (20.4–29.2) |  |
|  | > 9 members | 29.2 (20.2–38.2) |  |
| **Number of children under 5** |  |  | * |
|  | 0–1 | 20.5 (17.1–23.9) | 0.023* |
|  | 2–3 | 23.6 (19.4–27.8) |  |
|  | > 3 | 31.9 (22.3–41.4) |  |
| **Sex of household head** |  |  | ns |
|  | Male | 22.8 (18.6–26.9) | 0.827 |
|  | Female | 23.3 (19.6–27.0) |  |
| **Household wealth status** |  |  | * |
|  | Poorest | 35.2 (28.9–41.5) | <0.001* |
|  | Poorer | 33.4 (27.0–39.9) |  |
|  | Middle | 22.2 (16.5–27.9) |  |
|  | Richer | 10.7 (5.5–15.9) |  |
|  | Richest | 3.2 (0.5–5.9) |  |
| **Place of residence** |  |  | * |
|  | Urban | 9.7 (7.3–12.1) | <0.001* |
|  | Rural | 32.0 (26.9–37.1) |  |
| **Region** |  |  | * |
|  | Western | 31.4 (22.9–40.0) | <0.001* |
|  | Central | 29.3 (22.4–36.2) |  |
|  | Greater Accra | 1.0 (0.0–2.5) |  |
|  | Volta | 33.2 (17.1–49.2) |  |
|  | Eastern | 25.9 (15.9–35.8) |  |
|  | Ashanti | 15.8 (7.6–24.0) |  |
|  | Brong Ahafo | 35.7 (25.4–46.0) |  |
|  | Northern | 18.6 (12.2–25.1) |  |
|  | Upper East | 30.7 (22.6–38.8) |  |
|  | Upper West | 22.4 (15.3–29.5) |  |
| **Mosquito net ownership** |  |  | ns |
|  | No | 38.5 (–18.3–95.4) | 0.689 |
|  | Yes | 27.8 (23.7–31.9) |  |
| **Number of mosquito nets owned** |  |  | ns |
|  | 0 nets | 17.9 (12.2–23.6) | 0.356 |
|  | 1 net | 23.3 (17.0–29.6) |  |
|  | 2 nets | 24.1 (20.0–28.1) |  |
|  | 3 nets | 22.9 (17.1–28.7) |  |
|  | 4 nets | 23.0 (17.8–28.3) |  |
|  | 5 nets | 31.1 (19.9–42.3) |  |
|  | 6 nets | 19.5 (11.9–27.1) |  |
|  | 7+ nets | 24.0 (13.0–35.0) |  |
| **Household net usage (all/some/no child/no net)** |  |  | * |
|  | No | 14.7 (11.6–17.8) | <0.001* |
|  | All children | 27.7 (23.5–31.8) |  |
|  | Some children | 28.0 (20.6–35.3) |  |
|  | No net in household | 17.9 (12.2–23.6) |  |
| **Number of children under 5 who slept under a net** |  |  | * |
|  | No child | 15.9 (12.7–19.1) | <0.001* |
|  | 1–2 children | 27.5 (23.4–31.6) |  |
|  | > 2 children | 29.4 (20.0–38.8) |  |
| **Dwelling sprayed (12 months)** |  |  | Ns |
|  | No | 23.5 (19.9–27.1) | 0.151 |
|  | Yes | 18.5 (13.0–23.9) |  |
| **Household electricity** |  |  | * |
|  | No | 38.8 (33.2–44.3) | <0.001* |
|  | Yes | 18.6 (15.2–22.0) |  |
| **Household television** |  |  | * |
|  | No | 33.7 (29.0–38.4) | <0.001* |
|  | Yes | 16.4 (13.2–19.5) |  |
| **Child's age** |  |  | * |
|  | < 24 months | 16.7 (12.6–20.9) | <0.001* |
|  | 24–48 months | 24.9 (21.3–28.4) |  |
|  | > 48 months | 29.2 (23.8–34.6) |  |
| **Sex of child** |  |  | Ns |
|  | Male | 24.2 (20.1–28.2) | 0.118 |
|  | Female | 21.6 (18.4–24.9) |  |
| **Mother's education** |  |  | * |
|  | None | 29.4 (22.0–36.8) | <0.001* |
|  | Primary | 26.8 (21.1–32.6) |  |
|  | Secondary | 20.8 (16.9–24.8) |  |
|  | Bachelor's | 15.8 (9.5–22.1) |  |
|  | Higher | 0.3 (0.0–0.8) |  |
| **Child's anaemia level** |  |  | * |
|  | Not anemic | 13.1 (10.7–15.6) | <0.001* |
|  | Mild | 20.7 (16.5–24.9) |  |
|  | Moderate | 39.2 (33.6–44.8) |  |
|  | Severe | 72.1 (46.4–97.7) |  |

*where s denotes the set of sampled children, w□ the normalized sampling weight, and y□ the outcome (1 = positive malaria test, 0 = negative)*.

Design-based Rao–Scott chi-square tests corrected the standard Pearson statistic for the design effect of clustering and unequal weighting when testing bivariate associations between malaria status and each categorical covariate (Table 2).

The multivariate models in Table 3 were estimated as survey-weighted logistic regressions. For child i, let π□ denote the probability of a positive malaria test conditional on a vector of p covariates. The logit link function relating the log-odds of malaria positivity to the linear predictor is:

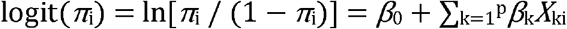

**Table 3.** Survey-weighted logistic regression models (quasibinomial link) of the determinants of malaria prevalence among children under five years. AOR = adjusted odds ratio; — = variable not included in that model; *p < 0.05. Source: Computed from GMIS 2019 using the R survey package.

| Variable | Model 1<br>AOR | Model 2<br>AOR | Model 3<br>AOR |
| --- | --- | --- | --- |
| Household size (ref: <6 members) |  |  |  |
| 6–9 members | 1.04 | 1.17 | 1.08 |
| >9 members | 1.32 | 1.52 | 1.48 |
| Place of residence (ref: Urban) |  |  |  |
| **Rural** | **2.02\*** | **2.17\*** | **2.36\*** |
| Household electricity (ref: Yes) |  |  |  |
| **No** | **1.50\*** | **1.38\*** | **1.17** |
| Region (ref: Greater Accra) |  |  |  |
| **Western** | **17.01\*** | **15.29\*** | **14.23\*** |
| **Central** | **15.87\*** | **18.20\*** | **14.80\*** |
| **Volta** | **12.51\*** | **12.35\*** | **11.44\*** |
| **Eastern** | **14.90\*** | **15.51\*** | **15.27\*** |
| **Ashanti** | **7.08\*** | **8.34\*** | **8.78\*** |
| **Brong Ahafo** | **13.26\*** | **13.91\*** | **11.71\*** |
| Northern | 3.63 | 2.70 | 1.69 |
| **Upper East** | **7.58\*** | **6.76\*** | **5.31\*** |
| Upper West | 4.84 | 5.06 | 5.79 |
| Sex of household head (ref: Male) |  |  |  |
| Female | 1.11 | 1.06 | 1.04 |
| Number of children under 5<br>(ref: 0–1) |  |  |  |
| 2–3 | 1.05 | 1.08 | 1.07 |
| >3 | 1.35 | 1.61 | 1.54 |
| Household wealth (ref:<br>Richest) |  |  |  |
| **Poorest** | **7.89\*** | **5.31\*** | **4.13\*** |
| **Poorer** | **6.66\*** | **4.14\*** | **3.13\*** |
| Middle | 4.22* | 2.74* | 2.00 |
| Richer | 2.53 | 2.02 | 1.57 |
| Mother's education (ref:<br>Higher) | — |  |  |
| **None** | — | **39.74\*** | **45.98\*** |
| **Primary** | — | **27.61\*** | **33.50\*** |
| **Secondary** | — | **25.06\*** | **30.28\*** |
| **Bachelor's** | — | **26.66\*** | **34.19\*** |
| Dwelling sprayed (ref: Yes) | — |  |  |
| **No** | — | **1.50** | **2.21\*** |
| Number of mosquito nets<br>(ref: 1 net) | — |  |  |
| 0 nets | — | 0.85 | 0.83 |
| **2 nets** | — | **0.67** | **0.61\*** |
| **3 nets** | — | **0.61\*** | **0.57\*** |
| **4 nets** | — | **0.49\*** | **0.47\*** |
| 5 nets | — | 0.92 | 1.07 |
| **6 nets** | — | **0.31\*** | **0.30\*** |
| 7+ nets | — | 0.74 | 0.81 |
| Children who slept under<br>net (ref: No child) | — |  |  |
| **1–2 children** | — | **1.24** | **1.48\*** |
| >2 children | — | 0.88 | 1.10 |
| Child's age (ref: <24<br>months) | — | — |  |
| **24–48 months** | — | — | **2.49\*** |
| **>48 months** | — | — | **4.17\*** |
| Sex of child (ref: Female) | — | — |  |
| Male | — | — | 1.14 |
| Child's anaemia level (ref:<br>Not anemic) | — | — |  |
| **Mild** | — | — | **1.74\*** |
| **Moderate** | — | — | **5.65\*** |
| **Severe** | — | — | **24.53\*** |

*where* β□ *is the intercept, X□□ is the value of the kth covariate for child i, and* β□ *is the corresponding log-odds coefficient, estimated by maximum quasi-likelihood under the survey design (cluster, strata, weight) described in Section 3.2*.

Adjusted odds ratios (AOR) reported in Table 3 were obtained by exponentiating the estimated coefficients, with 95% confidence intervals constructed from design-based standard errors:

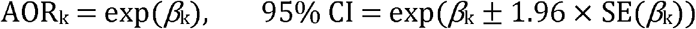

*where SE(*β*□) is the design-based (Taylor-linearized) standard error of the coefficient estimate*.

Two-way interaction terms (Section 4.4) added a product term X□□ × X□□ for covariates a and b to the linear predictor above, and each interaction’s joint contribution was assessed using a design-based Wald F-test comparing the model with and without the interaction term.

### 3.5 Limitations

Several limitations should be noted. First, the GMIS relies on self-reported data for several covariates, which is susceptible to social-desirability and recall bias. Second, as with all cross-sectional secondary data, the GMIS does not support causal inference between household, maternal, or preventive-behavior characteristics and malaria prevalence. Third, because several conceptually related covariates (e.g., wealth and electricity; number of nets owned and net usage) are correlated, coefficients for individual variables in the multivariate models should be interpreted jointly rather than in isolation, and the significant interaction effects reported in Section 4.4 indicate that several of these relationships are not adequately captured by a main-effects-only model. Fourth, and most importantly, anaemia is itself a recognized clinical consequence of acute and chronic malaria infection (through hemolysis and reduced red-cell production) rather than solely an independent risk factor for it. Including anaemia level as a predictor of malaria status in a cross-sectional model therefore risks conditioning on a variable that is downstream of, or bidirectionally related to, the outcome; the very large adjusted odds ratios observed for moderate and severe anaemia in this study (Table 3) most plausibly reflect this reverse or bidirectional relationship rather than anaemia acting as an upstream determinant of malaria risk. Readers should interpret the anaemia coefficients, and any resulting attenuation of other coefficients in the same model, with this important caveat in mind.

## 4. Results

### 4.1 Sample characteristics

Table 1 presents weighted characteristics of the 2,867 children under five years with a valid malaria RDT result. An estimated 22.9% (95% CI: 19.6–26.2%) tested positive for malaria. Most children (80.0%) were under 48 months old, and 59.5% lived in rural households. Household wealth was fairly evenly distributed, with the poorest quintile slightly overrepresented (24.1%) relative to the richest (14.8%). Nearly all households (99.8%) owned at least one insecticide-treated net, yet in 25.8% of households no child had slept under a net the previous night. Most mothers (71.4%) had at least a primary education, and most households had electricity (78.5%) and a television (62.2%). Just over half of children (54.8%) showed some degree of anaemia, including 1.4% with severe anaemia.

### 4.2 Bivariate associations

Table 2 presents weighted malaria prevalence by category, with Rao–Scott design-corrected chi-square tests of association for each characteristic. Household size, number of children under five, household wealth, place of residence, region, household net usage, number of children who slept under a net, household electricity, household television, child’s age, mother’s education, and child’s anaemia level were all significantly associated with malaria status (p < 0.05), with prevalence ranging from below 2% (Greater Accra region, mothers with higher education) to over 70% (children with severe anaemia). Sex of household head, mosquito net ownership, number of mosquito nets owned, dwelling spraying, and sex of child were not significantly associated with malaria status at the bivariate level.

### 4.3 Multivariate analysis

Table 3 presents adjusted odds ratios from the three survey-weighted logistic regression models. In Model 1 (household characteristics only), household wealth, place of residence, electricity, and region were significant predictors of malaria status: children in the poorest and poorer households had markedly higher odds of testing positive than children in the richest households (AOR = 7.89 and 6.66, respectively), rural children had roughly twice the odds of urban children (AOR = 2.02), and children in every region except Northern had significantly higher odds than children in Greater Accra, the region with the lowest observed prevalence.

Model 2 added mother’s education and preventive-behavior variables. Household wealth, residence, electricity, and region remained significant, and mother’s education emerged as a strong predictor: children of mothers with no more than a bachelor’s degree had 25-to 40-fold higher odds of testing positive than children of mothers with higher education (all p < 0.01). Owning three, four, or six mosquito nets was associated with significantly lower odds relative to owning one net, while dwelling spraying and household net-usage category were not significant.

Model 3, the final model, added child’s age, sex, and anaemia level. Household wealth (poorest and poorer quintiles), rural residence, region, and mother’s education all remained significant, though the electricity association attenuated to non-significance. Older children had significantly higher odds of testing positive than children under 24 months: AOR = 2.49 for children 24–48 months and AOR = 4.17 for children over 48 months. Anaemia level was strongly associated with malaria status, with adjusted odds ratios of 1.74, 5.65, and 24.53 for mild, moderate, and severe anaemia, respectively, relative to children who were not anemic — by far the largest associations observed in any model. Dwelling spraying became significant in this model (AOR = 2.21 for households not sprayed), and children in households where one to two children slept under a net had significantly higher, not lower, odds of testing positive than children in households where no child used a net (AOR = 1.48); this counter-intuitive direction is discussed further in Section 5.

### 4.4 Interaction effects

Because the conceptual framework suggests that maternal and household characteristics may act jointly rather than independently, two-way interaction terms were tested against the final model using design-based Wald F-tests (Table 4). The interaction between mother’s education and household wealth was highly significant (F = 128.03, df = 16, 124, p < 0.001), as was the interaction between mother’s education and children’s net usage (F = 12.50, df = 8, 132, p < 0.001). These results indicate that the association between mother’s education and malaria status varies substantially across household wealth levels and across household net-usage categories, and that the main-effects-only estimates for mother’s education in Table 3 should be interpreted as an average association that masks this underlying heterogeneity.

**Table 4.** Design-based Wald F-tests for two-way interaction terms added individually to the final multivariate model. *p < 0.05. Source: Computed from GMIS 2019 using the R survey package.

| Interaction term | F | df | p-value |
| --- | --- | --- | --- |
| **Mother's education × household wealth** | **128.03** | **16, 124** | **<0.001\*** |
| **Mother's education × children's net usage** | **12.50** | **8, 132** | **<0.001\*** |

## 5. Discussion

This study examined household, maternal, and child characteristics associated with malaria prevalence among children under five years in Ghana using a survey-weighted analysis of nationally representative data. Household wealth was a strong and consistent predictor across all three models, with children in poorer households having markedly higher odds of testing positive than children in the richest households. This gradient is consistent with the broader literature linking poverty to malaria risk through housing quality, environmental exposure, and access to preventive and curative care (Worrall et al., 2005; Somi et al., 2007; Bayode & Siegmund, 2022), and contrasts with weaker or null wealth effects reported in some smaller, non-survey-weighted Ghanaian samples (Filmer, 2005; Yusuf et al., 2010).

Region and place of residence remained significant net of wealth and other covariates, consistent with prior analyses of Ghanaian malaria data (Nyarko & Cobblah, 2014; Ejigu & Wencheko, 2021). Rural children had more than twice the odds of testing positive as urban children, and every region other than Northern had significantly higher odds than Greater Accra, likely reflecting differences in ecological and climatic conditions, vector density, and health-service coverage across the country.

Mother’s education was one of the strongest predictors of malaria status in this study, with children of mothers below the higher-education level facing dramatically greater odds of testing positive. This is consistent with evidence from Uganda (Masuda, 2020) and an earlier spatial analysis of Ghanaian GMIS data (Ejigu & Wencheko, 2021), and with the broader literature linking maternal education to child health outcomes through improved health-related knowledge, care-seeking behavior, and access to resources (Njau et al., 2014; Villamor et al., 2003). However, the significant interactions between mothers’ education and both household wealth and household net usage (Section 4.4) indicate that this protective association is not uniform: it is markedly stronger in some combinations of wealth and net usage than others, so the large main-effects odds ratios in Table 3 should not be read as applying equally across all households.

Older children had substantially higher odds of testing positive than children under 24 months, plausibly reflecting increasing exposure to mosquito bites as children become more mobile and spend more time outdoors, alongside a degree of acquired immunity that develops only gradually in this age range. The number of mosquito nets owned showed a partial protective pattern (households owning four or six nets had significantly lower odds than those owning one), but the household net-usage variables showed a less consistent and, for one category, counter-intuitive pattern: children in households where one to two children slept under a net had significantly higher odds of testing positive than children in households where no child used a net. This finding likely reflects reverse causation or residual confounding rather than nets increasing risk; households that have already recognized a child’s illness or a recent local outbreak may respond by increasing net usage, and net usage in a cross-sectional survey captures behavior on a single recent night rather than sustained practice. This finding should not be interpreted as evidence against the protective value of mosquito nets, which is well established in the experimental and quasi-experimental literature (Ong’echa et al., 2011; Somi et al., 2007; Ntonifor & Veyufambom, 2016).

Anaemia level showed by far the strongest association with malaria status of any variable in this study, with severe anaemia associated with roughly 25-fold higher odds of a positive malaria test. As noted in Section 3.5, this association should be interpreted with considerable caution: anaemia is a well-documented clinical consequence of malaria infection through parasite-induced hemolysis, and the cross-sectional design of this study cannot establish whether anaemia preceded, followed, or arose concurrently with the malaria episode captured by the RDT. The magnitude of this association, and the attenuation it produced in several other coefficients between Models 2 and 3 (notably electricity, which lost significance), suggests that anaemia may be functioning partly as a proxy for the outcome itself rather than as an independent upstream risk factor, and future analyses of these data may wish to model malaria and anaemia as related but distinct outcomes rather than treating one as a predictor of the other.

## 6. Conclusion and Recommendations

Household wealth, region, rural residence, mother’s education, child’s age, and child’s anaemia status were the principal correlates of malaria prevalence among children under five years in this nationally representative Ghanaian sample, and the significant interactions between mother’s education and both household wealth and household net usage indicate that maternal education’s protective association does not operate uniformly across the population. These findings support several policy directions. First, malaria-control programming should be explicitly pro-poor, since wealth showed a strong and consistent gradient across all models; poverty-reduction and social-protection measures may therefore have meaningful spillover benefits for child malaria risk. Second, health policymakers should sustain and expand efforts to improve women’s access to education, while recognizing that education-based behavior-change messaging may need to be tailored differently across wealth and net-usage strata to be equally effective. Third, targeted regional interventions are warranted in high-burden areas outside Greater Accra to identify the specific drivers of elevated malaria risk there. Finally, given the very strong association observed between anaemia and malaria status, integrated management of malaria and childhood anaemia rather than treating either condition in isolation may offer meaningful gains for child health in this population.

Given the cross-sectional and self-reported nature of the GMIS data, these findings should be interpreted as associations rather than causal relationships, and the anaemia and net-usage findings in particular warrant cautious interpretation for the reasons discussed in Sections 3.5 and 5. The significant interaction effects identified here also suggest that future analyses of these data should report and interpret stratified or interaction models rather than main-effects-only models, and future longitudinal research is recommended to clarify the direction and mechanisms underlying these relationships.

## Data Availability

The data used in this study are publicly available through the Demographic and Health Surveys (DHS) Program upon registration and approval of a data access request. https://dhsprogram.com/pubs/pdf/MIS35/MIS35.pdf
The R code used for data preparation and statistical analyses is available at https://github.com/Erico250/Malaria-Paper

https://github.com/Erico250/Malaria-Paper

## Acknowledgements

This manuscript is based on a dissertation submitted to the Regional Institute for Population Studies, University of Ghana, Legon, in partial fulfilment of the requirements for the degree of Master of Arts in Population Studies.

## Funding

No specific funding was received for this study.

## Data availability

The data used in this study are publicly available through the Demographic and Health Surveys (DHS) Program upon registration and approval of a data access request. The R code used for data preparation and statistical analyses is available at https://github.com/Erico250/Malaria-Paper

## Notes

### Competing Interest Statement

The authors have declared no competing interest.

### Author Declarations

The data used in this study are publicly available through the Demographic and Health Surveys (DHS) Program upon registration and approval of a data access request. https://dhsprogram.com/pubs/pdf/MIS35/MIS35.pdf The R code used for data preparation and statistical analyses is available at https://github.com/Erico250/Malaria-Paper

